# HRDetect in Ovarian Carcinoma: Stratification and Therapeutic Implications

**DOI:** 10.1101/2025.03.03.25323026

**Authors:** Kalyan Banda, Helen R Davies, Yogesh Kumar, Yasin Memari, Andrea Degasperi, Marc Radke, Isabel Rodriguez, Ted A. Gooley, Francesca Menghi, Thomas Harding, Kevin Lin, Edison T. Liu, Serena Nik-Zainal, Elizabeth M. Swisher

## Abstract

In this study, we show that comprehensive DNA profiling using whole genome sequencing (WGS) can offer novel insights into ovarian carcinoma (OC). First, using WGS of 185 OC with HRDetect profiling, a mutational signature-based algorithm for homologous recombination repair deficiency (HRD), we found that patients with HRDetect-high OC had significantly better overall survival, with a median of 6.2 years compared to 4.1 years for those with lower score OC. Second, the rest of the OCs are not simply non-HRD - some carry distinguishing rearrangement signatures that are strongly prognostic. Third, in an independent cohort of 77 patients, we demonstrate predictive value; HRDetect-high patients exhibited better responses to PARP inhibitors (PARPi) (median progression-free survival of 11.1 months versus 7.1 months; overall response rate of 54% versus 22.5%). Critical next steps include fine-tuning therapeutic vulnerabilities associated with the diverse rearrangement signatures, and harnessing the full potential of WGS for OC clinical trial stratification.

**Statement of Significance:** This study shows the HRDetect strongly predicts overall survival and PARPi response in ovarian carcinoma. Furthermore, we reveal that non-HRD tumors harbor distinct rearrangement signatures that confer significant prognostic value, underscoring the potential of WGS to refine patient stratification and guide tailored therapeutic strategies in OC.

## Introduction

In recent years, OC patients have benefited from the development of targeted therapeutics such as PARPi, which selectively kills homologous recombination repair deficiency (HRD) cancer cells. Yet, OC survival rates leave substantial room for improvement, as does our ability to precisely identify the right therapy for each patient. We investigated the potential of WGS approaches, including germline variation, somatic drivers, and mutational signatures for tailoring strategies in OC.

Mutational signatures are patterns of mutagenesis reporting endogenous and exogenous sources of DNA damage and repair, that have been operative throughout tumorigenesis. Through analytical, machine-learning, and experimental approaches, algorithms have been developed based on mutational signatures, aimed at improving clinical stratification of cancer genomes. HRDetect is an example, designed to detect *BRCA1*- and *BRCA2*-defective and other HRD cancers^1^. HRDetect leans on genomic patterns that are shared between *BRCA1-* and *BRCA2*-mutated cancers, including substitution signatures SBS3 and SBS8, microhomology-mediated deletions (mmdels), copy number-related global loss of heterogeneity (LOH), two distinguishing rearrangement signatures (RS) – RS3 enriched in *BRCA1*-mutated cancers, defined by tandem duplications (TDs) <10kb in length, and RS5 enriched in *BRCA2*-mutated lesions, defined by structural deletions <10kb in length. In triple-negative breast cancer (TNBC), HRDetect is an independent prognosticator amongst women receiving standard-of-care (SOC) treatment ^2^. Subsequently, HRDetect has been shown to predict a decline in circulating tumor DNA (ctDNA) in response to the PARPi rucaparib in treatment-naïve TNBC ^3^.

Building on the success of HRDetect in other tumor types, we sought to determine its prognostic value in OC. To this end, we performed WGS on matched tumor-normal samples from 185 patients with advanced-stage OC (the University of Washington (UW) cohort), a well-characterized group with extensive clinical follow-up. Further, to assess if HR detect correlates with PARPi sensitivity we performed WGS on match tumor normal samples from patients in ARIEL2, a one-arm phase II treatment trial of rucaparib in recurrent, platinum-sensitive, high-grade OC with measurable disease. Here, we report our findings and discuss their potential implications for refining therapeutic decision-making in OC

## Results

### Distribution and Implications of HRDetect in UW OC cohort

All patients in the cohort had undergone maximal effort surgical cytoreduction, and all but one received SOC chemotherapy with a platinum-doublet. Ten patients received neoadjuvant chemotherapy. WGS was performed on samples taken prior to treatment in 176 cases and from post-treatment recurrences in 9 samples (Supplementary data Fig1). Similar to TNBCs previously, more than half the cancers were HRDetect-high (95/185, 51.4% - exceeding a predefined score of 0.9, predictive of HRD), which correlated with a significantly improved overall survival (OS) (median OS 6.2 vs 4.1 years, HR=0.6, 95% CI=0.41-0.87, log-rank p=0.007) (Fig1a,h). A remarkable finding was that 23.2% (43/185) were HRDetect-intermediate (score 0.1-0.9), in contrast to only ∼13.5% of TNBCs^2^. Another 25.4% (47/185) were classified as HRDetect-low (score <0.1) (Fig 1a). Although HRDetect prognostically distinguished HRDetect-high from intermediate or low, (median OS 6.2 vs 4.8 vs 3.4 years; log-rank p = 0.02) (Fig 1i), there was no survival difference between individuals with HRDetect-intermediate and -low OC, out-of-keeping with TNBC where HRDetect-intermediate cases had the poorest outcomes^2^.

**Figure 1:**
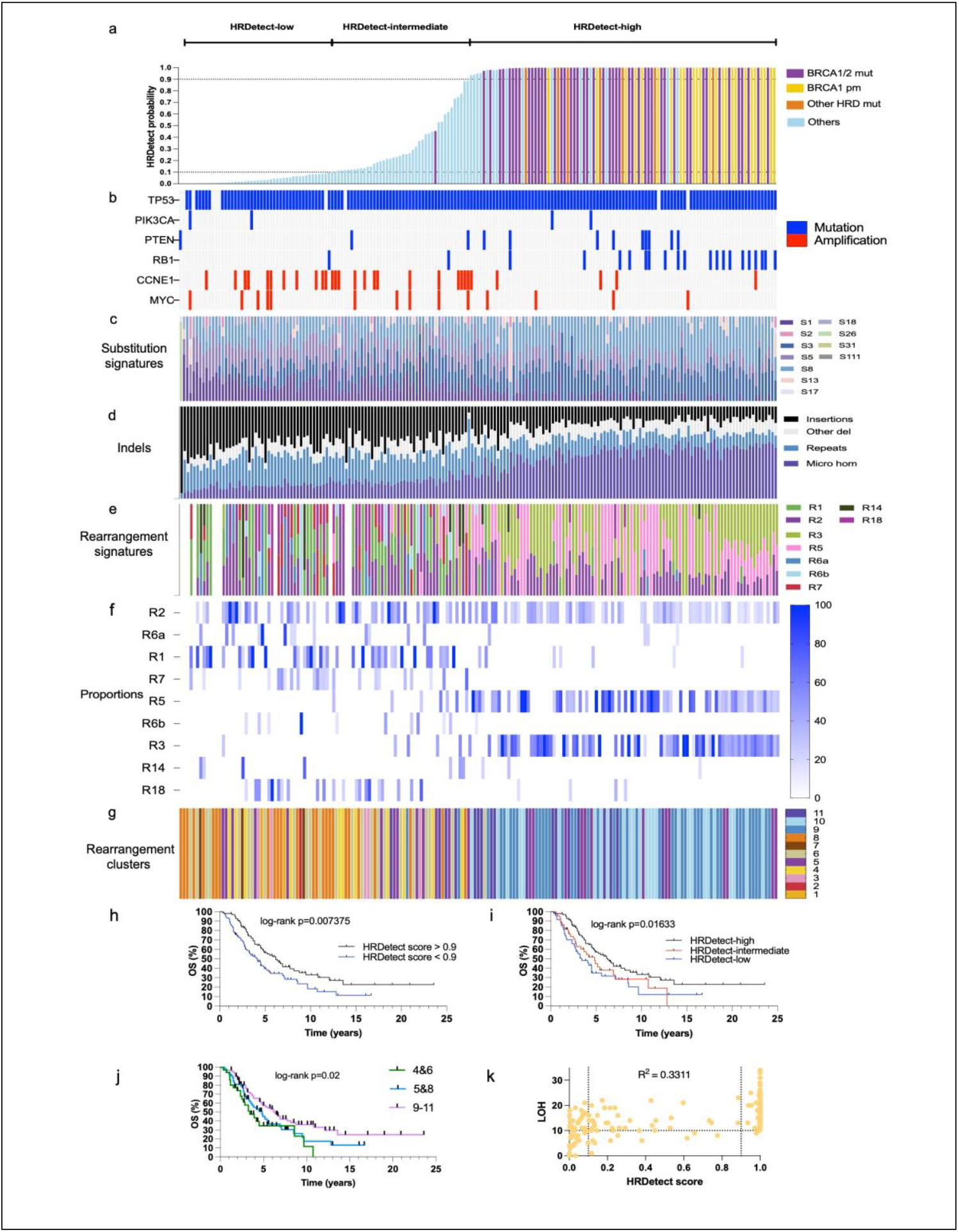
Clinical and genomic characteristics of HRDetect classification in the UW ovarian cancer cohort. A) Bar plot of HRDetect probability in 185 OCs b) genetic alterations in homologous recombination associated genes and other drivers c) d) & e) Proportions of mutational, indel patterns, and rearrangement signatures are shown as proportional bar plots, f) Proportions of rearrangement signatures as a heat map g) Rearrangement clusters of each case h) Kaplan–Meier analysis of association of OS with binary HRDetect classification i) Kaplan–Meier analysis of association of OS with HRDetect-high versus HRDetect-intermediate versus HRDetect-low j) Kaplan–Meier analysis of association of OS with rearrangement clusters 4&6 versus 5&8 versus 9-11 k) Correlation between genomic LOH and HRDetect scores.

### Genomic Features Associated with HRDetect Categories

We sought causes of high HRDetect scores. Of 95 HRDetect-high cases, 46 (48.4%) had deleterious germline and/or somatic mutations in *BRCA1* or *BRCA2* with concomitant loss of the wildtype allele (Fig 1a and Supplementary data Fig 5). Promoter hypermethylation was observed in *BRCA1* in 21 (22.1%) and in *RAD51C* in one (1%). Pathogenic germline variants were observed in other key HRD genes in 6 cases (6.3%); 2 in *RAD51C*, one in *RAD51D*, one in *PALB2*, and two in *BRIP1*, all with loss of the other allele. The causes for HRDetect-high scores in the residual 21 (22.1%) remain unexplained, although 4 OC had *BRCA1* promoter hypermethylation with LOH, albeit below the predefined quantitative cut-off of 60% methylation^4^. Notably, these 21 HRDetect-high cases would have been missed using conventional targeted-sequencing panels and gene-specific epigenetic assays. While the application of customary copy-number-based gLOH assay obtained from WGS would have captured 93 of 95 cases, it will also have called an additional 62 of 90 remaining cases as HRD, suggesting lower specificity for gLOH(Fig1k)^5^.

We also asked whether there were prognostically differentiating driver mutations in the UW cohort (Fig1b, Supplementary data Fig 3b). 19/21 (90.4%) of the cases had which *RB1* driver mutations were enriched amongst HRDetect-high platinum-sensitive cases (p=0.0005). Interestingly, the singular *RB1* mutant case that had platinum-refractory disease (progression during primary adjuvant chemotherapy) had a HRDetect-low score. 29 OC had *CCNE1* amplifications, of which two were platinum-refractory. *CCNE1* amplifications were underrepresented in HRDetect-high cases and equally frequent in HRDetect-intermediate and -low categories. Each had another driver event - a *MYC* amplification and a *CDK12* mutation respectively. All other drivers were not adequately powered for a clear analysis but were without clear prognostic impact in this cohort. 14 OC had *MYC* amplifications and distributed across all three HRDetect categories with 4, 5, and 5 cases in the high, intermediate, and low respectively. Two of these cases were platinum-refractory and had another driver event – a *PIK3CA* mutation and the *CCNE1* amp mentioned above. Four OC had *CDK12* drivers. One was HRDetect-high and platinum-sensitive, two were HRDetect-intermediate, one platinum-sensitive, and one platinum-refractory, and one was HRDetect low with unknown platinum status. There were also four OC with *PIK3CA* mutations (two HRDetect-high and two low).

### Rearrangement Signatures as Classifiers and Prognosticators Beyond HRDetect

In clinical settings, OC is often dichotomously defined as HRD or HR proficient (HRP). Critically, we find that the ‘HRP’ category is highly heterogeneous, enriched with distinguishing rearrangement signatures (RS) including RS1, RS14 (long tandem duplicator phenotypes), R2, R6a, and R6b (characterized by complex, focal amplifications), and R18 (defined by inter-chromosomal translocations) (Fig 1 a,e,f, Supplementary data Fig 3). To independently evaluate the importance of these RS, we performed unsupervised hierarchical clustering using only RS and evaluated the OS of resulting clusters. Eleven clusters were identified (Fig 1 g, Supplementary data Fig 4). Focusing on clusters that had greater than 10 samples, clusters associated with HRD (clusters 9, 10, and 11) were characterized by RS3 and RS5 and predominantly had very high HRDetect scores (74/87 >= 0.98)). They mirrored HRDetect prognostication and most had favorable outcomes (Figure 1j). Cluster 4, characterized by a high proportion of RS2, and cluster 6, with high proportion of RS1, were associated with the poorest outcomes, while the intermediate outcome clusters contained those with relatively quiet genomes (cluster 8) and cluster 5, containing a mixture of samples with both RS2 and RS3. These results suggest that in the HRP category, rearrangement signatures may serve as discriminating biological signals, and can make a further contribution towards prognostication, beyond HRDetect scoring alone.

### Characterizing HRDetect-Intermediate and -Low OC

To better understand this, we evaluated the 90 HRP cases in more detail. First there were commonalities between the 43 HRDetect-intermediate (probability score between 0.1 and 0.9) and 47 HRDetect-low cases (score <0.1): 30 of 43 (69.8%) and 30 of 47 (63.8%) cases of HRDetect-intermediate and -low respectively, carried long TD phenotypes of RS1 or RS14 and/or a substantial contribution from RS18 (translocations), compared to 14/95 (14.7%) of HRD (HRDetect-high (score >0.9)). The strongly negative prognostic TD phenotypes are prevalent in both intermediate and low groups, hence, survival analyses contrasting HRDetect-intermediate and -low are non-discriminating. Of the remaining 13/43 HRDetect-intermediate cases, 10 had R2-related translocations or R7-long deletions (>10kb). Two were chromosomally quiet genomes. Amongst HRDetect-low cases, of the remaining 17/47 cases, 8 had RS2, RS6a or 6b. Nine were chromosomally quiescent, and one case had a striking mismatch repair deficiency, with the typical substitution (SBS26 characterized by T>C transitions) and indel features (small insertions at poly nucleotide repeat tracts) caused by *PMS2* abrogation, a known cause of mismatch repair deficiency as confirmed through both cancer analyses ^6^and in human experimental systems ^7^.

### Predictive Value of HRDetect in the ARIEL2 Cohort

To next assess whether HRDetect could be predictive of sensitivity to PARPi, we performed WGS in paired tumor-normal DNA from ARIEL2, a one-arm phase II treatment trial of rucaparib in recurrent, platinum-sensitive, high-grade OC with measurable disease. 77 cancers were evaluated by WGS. 37(48%) were HRDetect-high, 12(15%) HRDetect-intermediate, and 28(37%) were HRDetect-low (Fig 2a). The proportions in the ARIEL2 cohort are different from the UW cohort, reflecting differential selection for relapsed, platinum-sensitive OC in the context of clinical trial.

**Figure 2:**
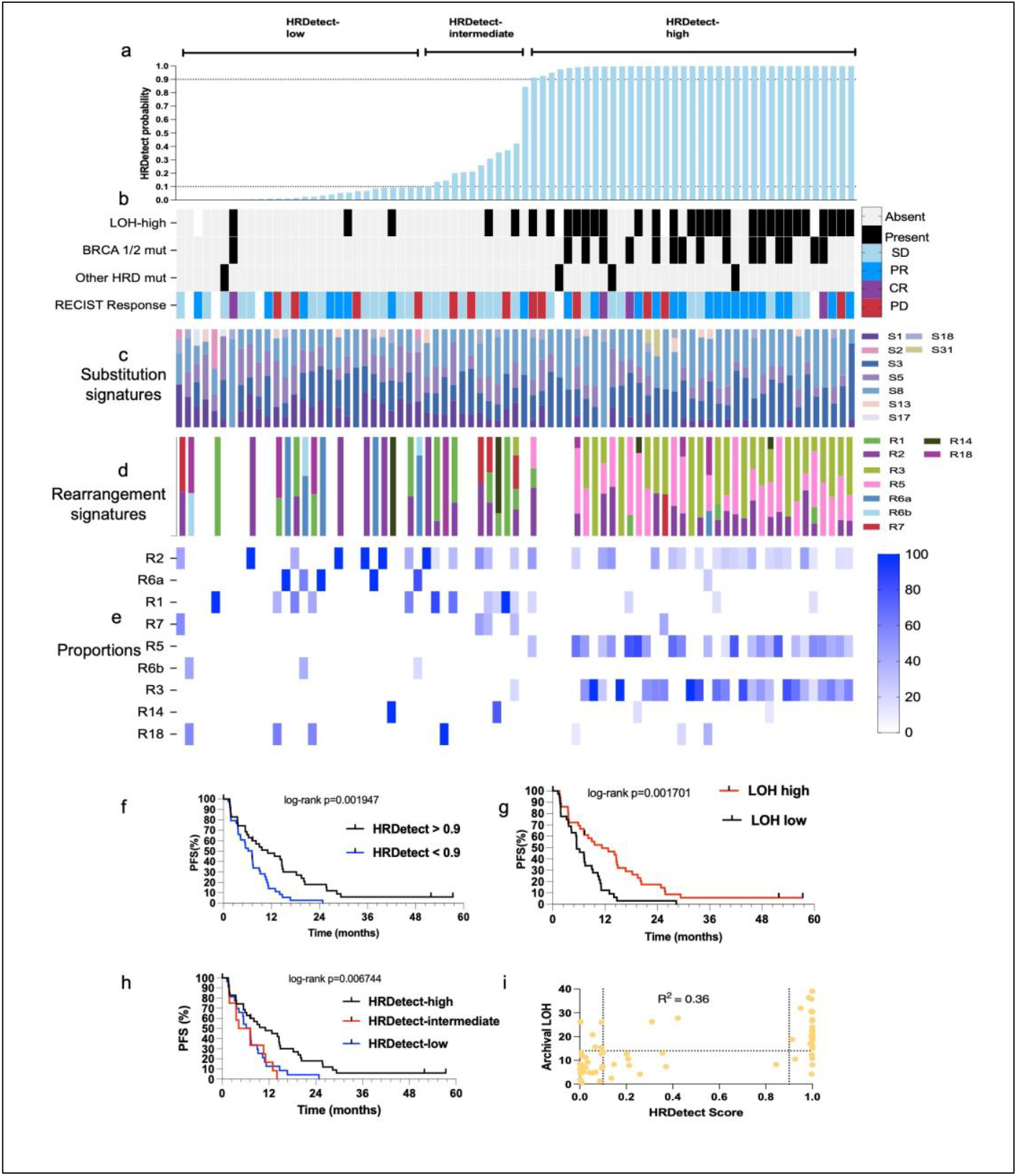
Clinical and genomic characteristics of HRDetect classification in patients treated with rucaparib in the ARIEL 2 clinical trial. a) Bar plot of HRDetect probability in 77 OCs on clinical trial b) genetic alterations in homologous recombination associated genes and other drivers, clinical responses to PARPi therapy c) & d) Proportions of mutational and rearrangement signatures are shown as proportional bar plots, e) Proportions of rearrangement signatures as a heat map f) Kaplan–Meier analysis of association of PFS with binary HRDetect classification g) Kaplan–Meier analysis of association of PFS with genomic LOH h) Kaplan– Meier analysis of association of PFS with HRDetect-high versus HRDetect-intermediate versus HRDetect-low i) Correlation of genomic LOH with HRDetect scores.

Patients with HRDetect-high OC treated with PARPi had a significantly improved progression-free survival (PFS) compared to the rest (median PFS 11.1 vs 7.1 months, HR=0.44, 95% CI=0.26-0.74, log-rank p=0.03) (Fig 2f) and treatment overall response-rates (ORR=54% vs 22.5%), demonstrating that HRDetect is an excellent identifier of HRD and predictor of therapeutic response to PARPi in OC. Causes of HRD were identified in 56.8% of HRDetect-high cases including 15 (40.5%) with germline or somatic mutations in *BRCA1/BRCA2*, one germline mutation in each of *RAD51C, BRIP1*, and *RAD51D*, and three with *BRCA1* promoter hypermethylation (Fig 2b). The causes for HRDetect-high scores in the remaining 16 (43.4%) samples were unclear. Enrollment criteria for ARIEL2 enriched for BRCA wildtype cases ^8^, explaining the higher fraction of unexplained high HRDetect in this cohort.

### Comparison with Genomic LOH HRD and Implications for Clinical Use

Finally, we compared with genomic LOH HRD as defined by FoundationOne™ NGS assay used in ARIEL2. There was a modest correlation R^2^ = 0.34 (Fig 2i) with 29 (78.4%) HRDetect-high cases also having high LOH. 7 (17.5%) of HRDetect-low/intermediate had high LOH. 6 cases were called LOH high but did not have a high HRDetect score nor carried mutations to explain reported LOH. Median PFS for LOH high vs low cases was 11.3 vs 5.5 months (HR= 0.49, 95% CI = 0.29-0.87, log-rank p=0.0017) (Fig 2g) as a predictor on PARPi therapy, contrasting the curves between HRDetect and LOH did not reveal a significant difference between them.

In summary, our study demonstrates that HRDetect prognosticates effectively and accurately predicts HRD and PARPi response in OC. Beyond HRDetect, WGS reveals a continuum of biology and survival in non-HRD OC. Key indicators of improved response to SOC include a very high HRDetect score (>0.98) and *RB1* mutations. Of other HRDetect groups, *CCNE1* amplifications could help improve prognostic status. However, high RS1, RS14, and RS18 are likely indicators of poor outcomes and alternative therapeutic strategies must be considered for these patients. Crucially we now have a way of stratifying these cancers using WGS strategies.

## Discussion

Our study definitively demonstrates that HRDetect and rearrangement signatures stratify OC beyond the binary HRD/HRP classification and reveal a continuum of genomic complexity. Unlike endometriosis-related cancers (clear cell and endometrioid), high-grade serous cancers (HGSOC) have not been stratified primarily by single nucleotide variant mutational signatures^9^. Large-structural features such as foldback inversions (FBI) with high-level amplifications are reported as a dominant prognostic feature and classify HGSOC into two H-FBI and, H-HRD (with duplications or deletion rearrangements indicative of HRD). Other studies have shown a high prevalence of *BRCA*-associated and age-associated-signatures, and a small number of cases with APOBEC or mismatch repair signatures ^10^. These studies in essence have still created a binary classification. Shallow WGS studies of copy number signatures have shown a more nuanced picture with seven signatures associated with survival and separated into three clusters that did not provide additional prognostic information^11^. Our data which primarily included HGSOC significantly expands on this. Not only are individual rearrangement signatures such as RS1 and RS1 linked to poorer prognoses but more importantly, unsupervised clustering revealed eleven RS-defined clusters that refine prognostication further and suggest that there is significantly more genomic heterogeneity in OC than previously thought. Understanding this heterogeneity is particularly important for the nominally homogenous HRP category which has worse outcomes, reduced benefit from PARPi, and requires distinct therapeutic strategies. Exciting future opportunities include mapping the immunophenotypic and therapeutic landscape of these RS-defined clusters.

HRDetect, initially developed and validated in breast cancer ^1 2^, is a robust marker of HRD in OC, not only identifying cancers with canonical HRD features but revealing a substantial fraction without overt *BRCA-*alterations. HRDetect-high OC were associated with a significantly improved overall survival confirming that the prognostic implications in OC, similar to TNBC. A striking difference is the prevalence of HRDetect-intermediate category in OC, constituting 25% of all cases and 50% of HRP, compared to ∼ 13.5% of all TNBC. Furthermore, the HRDetect-intermediate had an “intermediate” overall survival compared to the HRDetect-high and low categories, in contrast to its poor prognosis in TNBC. These findings suggest that HRDetect-intermediate represents a different biology in OC than TNBC, and presents another opportunity to prognosticate and target a large subset of OC. As sequencing costs decline and computational methods evolve, WGS approaches will be more readily accessible globally. Integrating HRDetect and rearrangement signatures into prospective clinical trials and routine practice could refine patient stratification.

HRDetect-intermediate and low categories could not be distinguished by driver mutations confirming that WGS can provide a refined view of heterogeneity that cannot be captured by targeted sequencing. Both had an equal distribution of *CCNE1* amplifications, with a few cases in the HRDetect-high group. *CCNE1* amplifications are thought to be mutually exclusive with *BRCA* alterations mutations^12^, are generally associated with unfavorable outcomes ^13,14^, and are of great interest as a therapeutic target ^15 16^. Our data suggest that *CCNE1* amplified OC may be composed of distinct groups with various mutator signatures. In this context, WGS and rearrangement signatures could also be viewed as important research tools to help understand the pathogenesis and delineate the heterogeneity even within OC with hallmark drivers (*CCNE1, CDK 12*, etc).

Currently utilized biomarkers of HRD are instrumental in guiding therapy with PARPi, but they capture only part of the underlying biological complexity, leading to variable response rates ^17 18^. Since HRDetect synthesizes multiple genomic features into a single score, it may better identify the underlying DNA repair defect than any individual marker. This expanded view of HRD is clinically relevant and can extend the benefits of PARPi. Patients with HRDetect-high OCs responded significantly better to rucaparib, suggesting that HRDetect may be a predictive biomarker for PARPi sensitivity, as previously speculated in TNBC ^3^. However, the limited number of samples available from ARIEL2 left us underpowered to compare with the commercially available HRD test based only on genomic LOH. Therefore, larger studies with well-defined cohorts treated with a PARPi are needed to confirm HRDetect as a predictive marker of PARPi sensitivity.

In conclusion, our study places HRDetect and rearrangement signatures into the broader context of OC classification, and treatment. It highlights the complexity of cancer biology that lies beyond traditional biomarkers. By synthesizing HRDetect and RS analysis, we can open the door to more finely tuned clinical strategies in a disease that remains a leading cause of gynecologic cancer mortality.

## Methods

### Ethics approval and consent to participate

Ethics approval and consent to participate. The UW Ovarian Cancer Biorepository was approved by the Institutional Review Board (IRB). All patients provided written informed consent prior to enrolment. Likewise, ARIEL2 participants provided informed consent, including for translational studies.

### University of Washington (UW) Patient cohort

Patients diagnosed with high-grade OC, undergoing surgery, at UW were prospectively enrolled in a biorepository. Of these, 212 representative patients with 222 samples were selected for this study (Supplementary fig 1). 31 samples were eliminated from analysis for quality control reasons leaving 191 neoplastic samples from 185 patients. There were 170 (92%) patients with samples collected at primary debulking surgery only, 9 (5%) patients with samples collected at recurrence only, and 6 patients (3%, 12 samples) with samples collected at both presentation and recurrence. All patients were followed until death and, if alive, censored at the last follow-up.

### ARIEL2 cohort

ARIEL2 part 1 was an international, multicenter, phase 2, open-label study of oral rucaparib in patients with recurrent, platinum-sensitive, high-grade ovarian carcinoma, which were classified into one of three predefined homologous recombination deficiency subgroups based on tumor mutational analysis: *BRCA* mutant (deleterious germline or somatic), *BRCA* wild type and LOH-high (LOH-high group), or *BRCA* wild type and LOH-low (LOH-low group). A cutoff of 14% or more genomic LOH was prespecified for the LOH-high group ^8^.

### Tissue sampling, DNA, and RNA extraction

Fresh tumor samples preserved in RNAlater (Qiagen) were obtained with routine clinical sampling. RNA and DNA were extracted using the Qiagen Allprep extraction kit (Qiagen) as previously described. ARIEL2 included DNA extraction formalin fixed paraffin tissues (FFPE) as previously described ^19^.

### Whole genome sequencing analysis

DNA extracted from both the tumor and corresponding normal tissue were subjected to whole-genome sequencing to an average sequencing depth of 29X in the tumor and 27X in the matched normal on Illumina sequencers. The resulting BAM files were aligned to the reference human genome (GRCh37) using Burrows-Wheeler aligner, bwa mem 0.7.17-r1188 (dockstore-cgpmap 3.0.4 https://quay.io/repository/wtsicgp/dockstore-cgpmap. Mutation calling was performed as described previously^10,11 .19,^The algorithms used are contained in the following dockstore,(dockstore-cgpwgs v2.1.0 https://quay.io/repository/wtsicgp/dockstore-cgpwgs).

### Mutational signatures and HRDetect

Mutational signatures were assigned using FitMS 2.4.1 ^21^. For substitution signatures, a two-step approach was used, first fitting a set of reference signatures commonly found in ovarian cancer, followed by rare signatures previously found in ovarian cancer. This two-step approach improves the solution in cases where substantial residual mutations remain after fitting the common signatures, whilst minimizing inappropriate overfitting of mutational signatures. HRDetect is a weighted model that combines the contributions of six distinguishing mutational signatures to produce a signal score that is predictive of a defect in the homologous recombination DNA repair pathway^1^. Following signature assignment, the contributions of substitutions signatures SBS3 and SBS8, rearrangement signatures RefSig R3 and RefSig R5, HRD LOH index and proportion of small deletions at regions of microhomology were used to calculate the HRDetect probability score. Code for the versions of both FitMS and HRDetect used can be found here (https://github.com/Nik-Zainal-Group/signature.tools.lib.)

### Hierarchical clustering

Column-based hierarchical clustering of rearrangement reference signatures was performed with a default setting of pheatmap package (Version 1.0.12) and cutree_cols setting was set as 10. Visualization plots were created with ggplot2(2.2.1) and custom R scripts.

### Methylation analyses

Neoplastic DNA was bisulfite converted using the EZ DNA Methylation-Lightning kit (Zymo Research). Droplet digital PCR was performed using the QX200 AutoDG System (Bio-rad laboratories). Primers were designed targeting a 72-bp amplicon in the *BRCA1* promoter region associated with transcriptional repression, with specific QSY hybridization probes targeting the fully methylated (FAM-labeled) and the fully unmethylated sequences (VIC-labeled). Quantitative methylation (fraction of methylated droplets divided by all positive droplets) was corrected for neoplastic cellularity from whole genome sequencing estimation. Samples with ≥ 60% methylation were considered positive for methylation.

Methylation analysis on samples from ARIEL2 was done as previously described ^22^

### Statistical analyses

Survival analyses and figures were created in GraphPad Pris,(version 10.0.2). Survival curves were compared using Kaplan–Meier estimates and the log-rank test. Hazard ratios were calculated through univariable or multivariable Cox regression using the coxph R function. Statistical comparisons between groups were performed using Wilcoxon’s or Kruskal–Wallis tests for numerical values or the chi-squared test for ordinal values. All P values reported from statistical tests are two-sided if not otherwise specified.

## Data Availability

All data produced in the present study are available upon reasonable request to the authors

https://quay.io/repository/wtsicgp/dockstore-cgpmap

## Acknowledgments

We gratefully acknowledge the support for K.B.’s research from the American Society of Clinical Oncology/Conquer Cancer Foundation’s Career Development Grant and the generosity of Roger Wilcox. The research in E.S.’s laboratory received funding from the Department of Defense Ovarian Cancer Research Program Clinical Development Award (OC160274). Additionally, this research was supported by Seattle Translational Tumor Research (STTR). The work in S.N.-Z.’s laboratory was made possible through funding from several prestigious sources: Cancer Research UK (CRUK) Advanced Clinician Scientist Award (C60100/A23916), the Dr. Josef Steiner Cancer Research Award 2019, the Basser Gray Prime Award 2020, the CRUK Grand Challenge Award (C60100/A25274), and the National Institute of Health Research (NIHR) Research Professorship (NIHR301627). Support was also provided by the NIHR Cambridge Biomedical Research Centre (BRC-1215-20014). The opinions expressed in this publication are those of the authors and do not necessarily reflect the views of the NIHR or the Department of Health and Social Care.

## Supplementary Data Figures

**Supplementary Figure 1:**
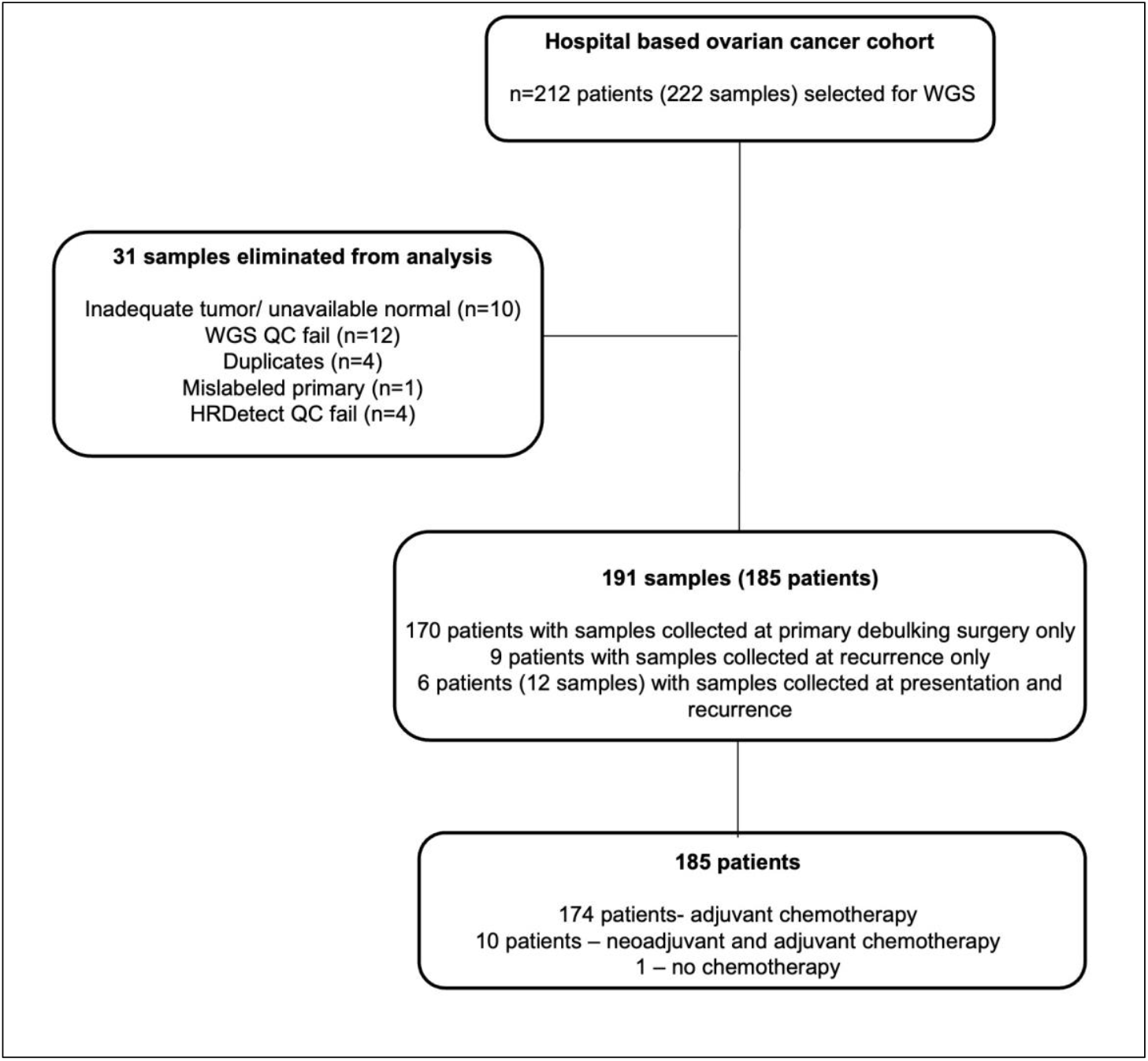
CONSORT diagram of the UW ovarian cancer cohort.

**Supplementary Figure 2:**
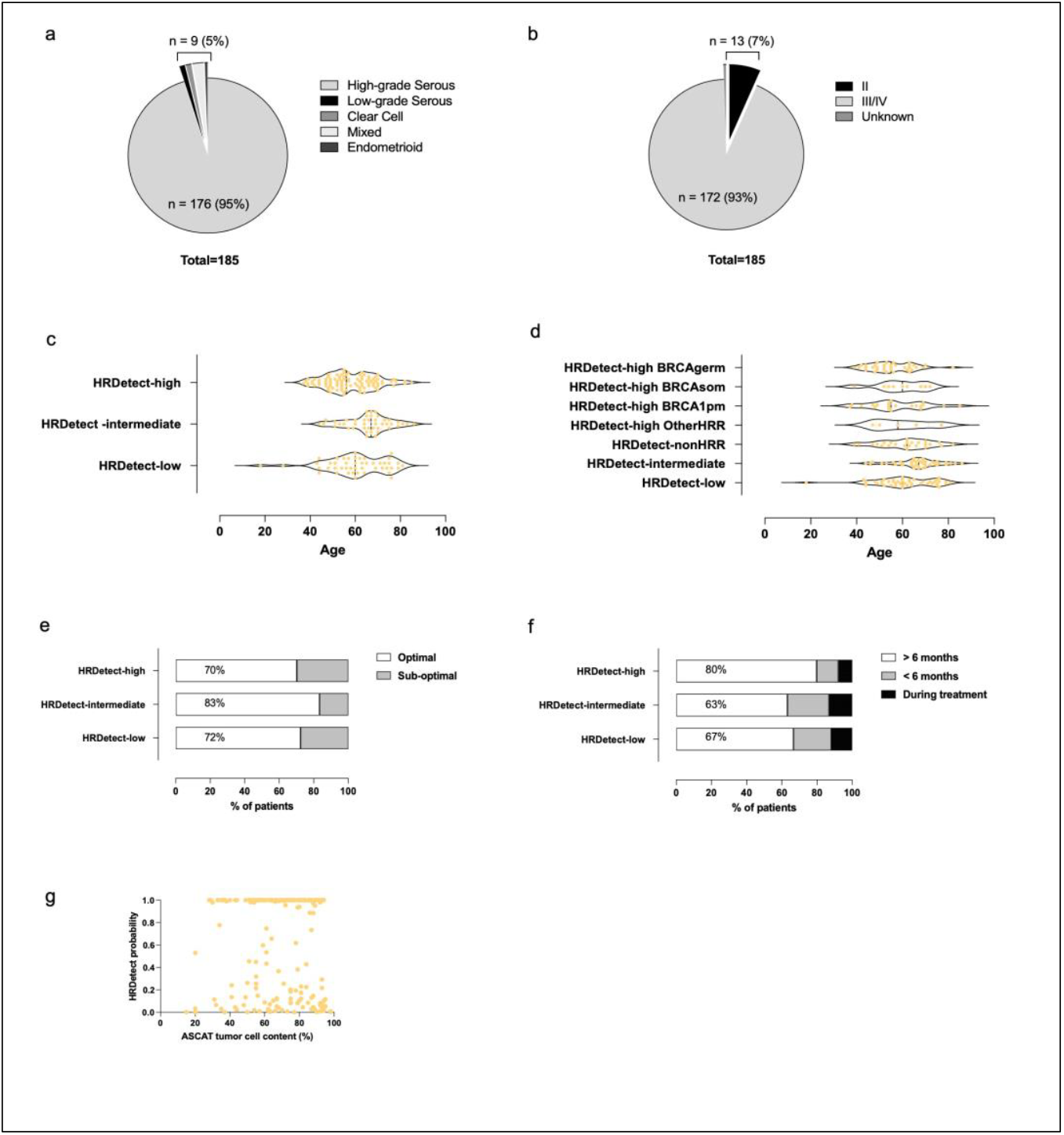
Clinico-pathological characteristics of HRDetect classification in the UW ovarian cancer cohort. A) and b) histology and stage. Age distribution by c) HRDetect classification d) According to drivers of HRDetect-high e) Surgical outcomes f) rate of recurrence g) correlation between ASCAT tumor percentage and HRDetect

**Supplementary Figure 3:**
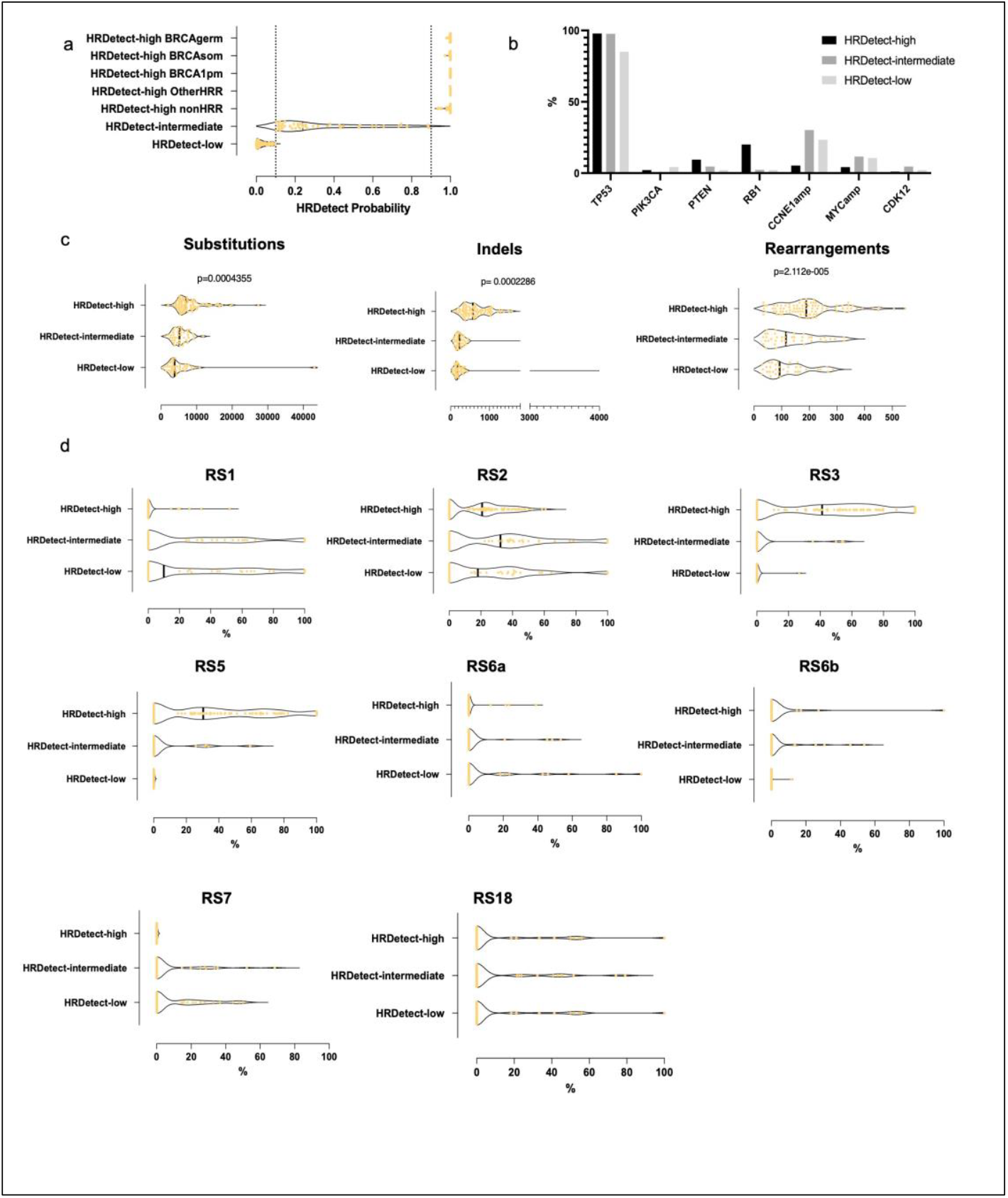
Genomic characteristics in HRDetect groups. a) Distribution of HRDetect scores b) Key driver alterations c) Number of Indels, substitutions, and rearrangements d) Proportions of individual rearrangement signatures.

**Supplementary Figure 4:**
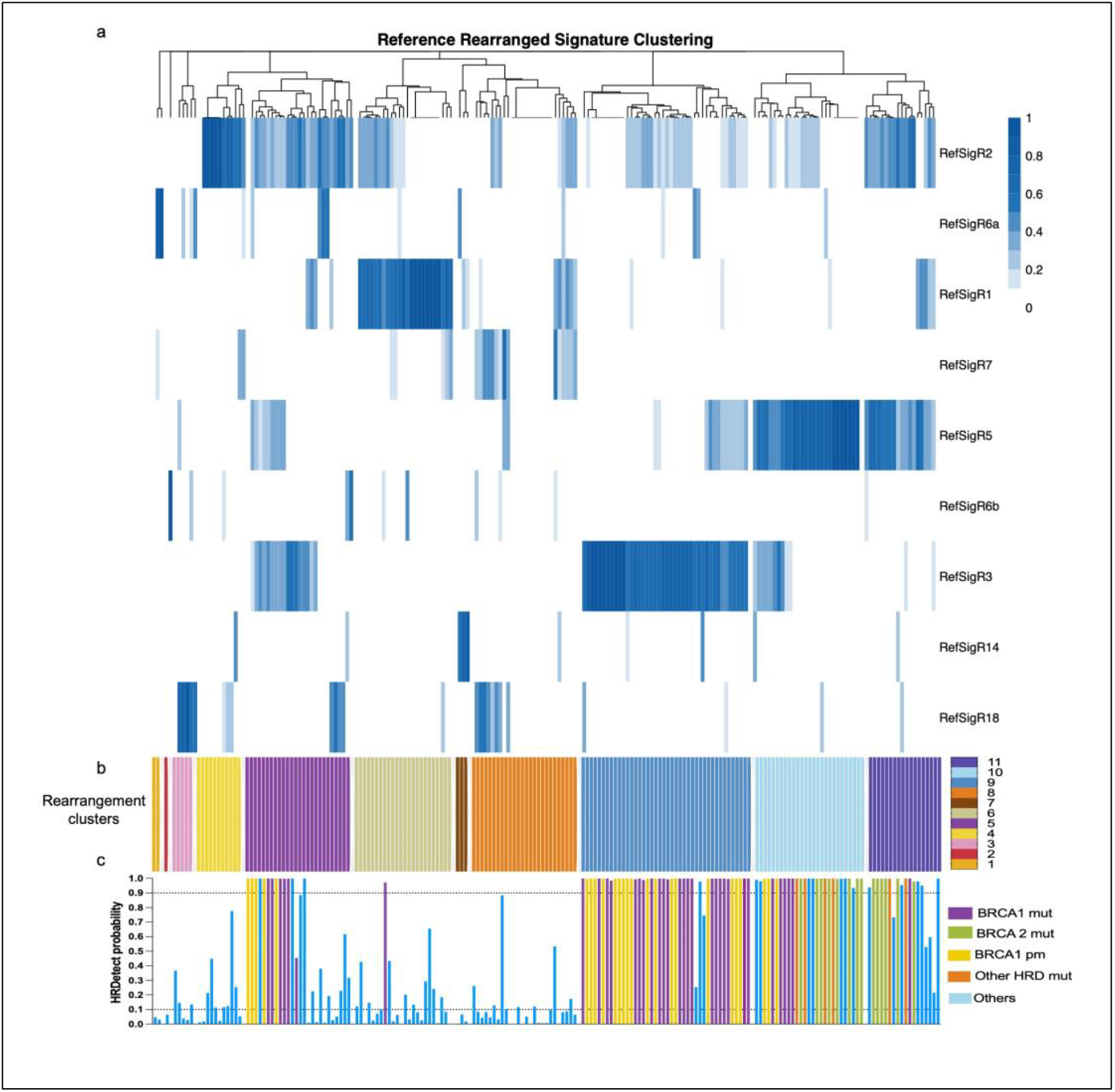
Hierarchical clustering by rearrangement signatures. a) Dendrogram and heatmap of proportions b) rearrangement clusters c) HRDetect probability

**Supplementary Figure 5:**
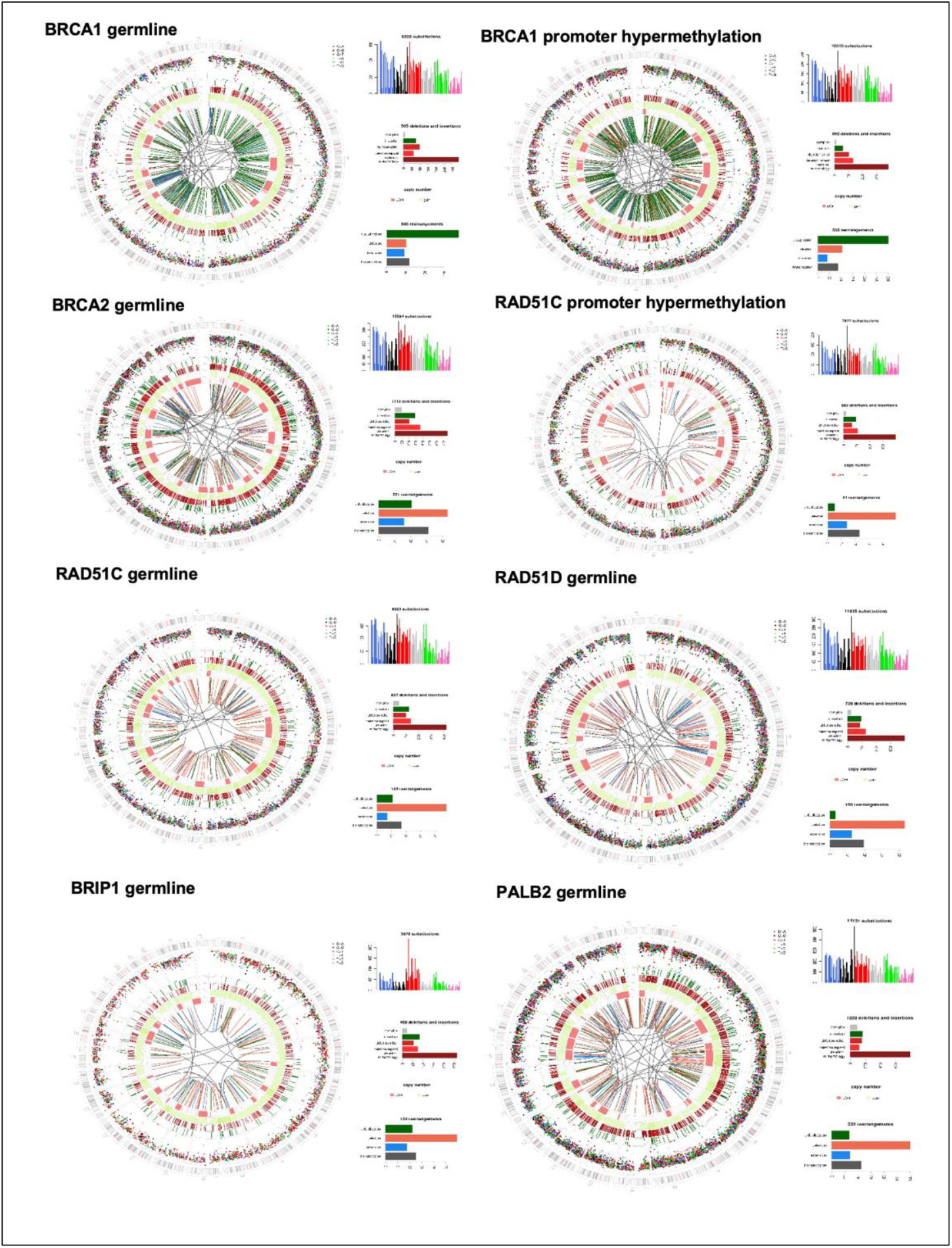
Genomic characteristics of HRDetect-high cases with known HRR mutations.

